# Lessons from a pandemic

**DOI:** 10.1101/2022.03.30.22273190

**Authors:** Yves Eggli, Valentin Rousson

## Abstract

**Objectives:** Several interventions have been used around the world trying to contain the SARS-Cov-2 pandemic, such as quarantine, prohibition of mass demonstrations, isolation of sick people, tracing of virus carriers, semi-containment, promotion of barrier gestures, development of rapid auto-tests and vaccines among others. We propose a simple model to evaluate the potential impact of such interventions.

**Methods:** A model for the reproduction number of an infectious disease including three main contexts of infection (indoor mass events, public indoor activities and household) and seven parameters is considered. We illustrate how these parameters could be obtained from the literature or from expert assumptions, and we apply the model to describe 20 scenarios that can typically occur during the different phases of a pandemic.

**Results:** This model provides a useful framework for better understanding and communicating the effects of different (combinations of) possible interventions, while encouraging constant updating of expert assumptions to better match reality.

**Conclusion:** This simple approach will bring more transparency and public support to help governments to think, decide, evaluate and adjust what to do during a pandemic.

## Introduction

The SARS-Cov-2 pandemic has been a challenge for the whole world. Countries implemented diverse measures to contain it such as quarantine, prohibition of mass demonstrations, isolation of sick people, tracing of virus carriers, semi-containment, promotion of barrier gestures (masks, hand washing with alcohol), development of rapid auto-tests and vaccines among others. The effectiveness of such specific interventions is difficult to measure, since they were often implemented simultaneously and in non-comparable contexts [1].

Politicians often turned to scientists with the creation of “task forces” to advise them. They took tough political decisions, depriving residents of their freedom of movement, shutting down schools and entire sectors of the economy for several months [2]. Fear set in and conflicting information circulated [3]. They invested significant amounts in scientific research [4]. Controversies arose, for example on the effectiveness of certain treatments, on the importance of the climatic factor [5, 6] or on the impact of partial containment measures [7, 8]. This planetary experience showed that it was difficult to predict the development of a pandemic [9, 10]. How should we react if a new pandemic would occur? What lessons can we learn from the current pandemic?

From our point of view, the main lesson is that we missed a general theory to manage this pandemic, in a context of great uncertainty. We missed a conceptual framework to assess the potential impact of each intervention, taking into account the characteristics of the virus and the affected population.

The aim of this paper is to lay the foundations for such a theory for the different phases of a pandemic. Most of the elements of this theory are known and have already been intuitively applied. The purpose here is only to clarify certain points and to set the minimum parameters necessary to monitor and manage a pandemic. Everyone can then refine the proposed model and make their own assumptions and adjustments.

## Methods

A formula has long been available for measuring the development of an epidemic through its reproduction number R, defined as the average number of people infected by a carrier of a specific infection [11,12]. If the value of R exceeds 1.0 the epidemic will grow up exponentially, if it is less than 1.0 it will die out. This formula involves the product of three factors as follows:

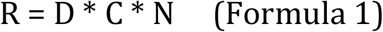

where D is the average duration of contagion (measured in days), C is the contagiousness (probability that a person will be contaminated during a contact with a carrier), and N is the average number of contacts per day that a carrier may have. Of course, the contagiousness will not be the same for each contact, obviously depending on the “intensity” of the contact. In this formula, C must therefore be considered as an “average” probability calculated over all types of daily contacts.

Formula 1 applies at the start of an epidemic, where it is usually noted R_0_, and when anyone in contact is likely to be infected. To take into account the appearance of immunized people (following the disease or by vaccination [13]), a fourth factor is introduced into the formula [14], yielding:

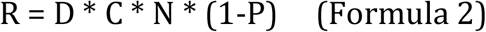

where P is the prevalence of immunized people. Formula 2 explains herd immunity, the value of R falling below 1.0 if P is large enough, in fact as soon as P>1-1/R_0_.

From there, one can immediately understand how the various measures taken by a government may affect the course of an epidemic. Testing and isolating the patients makes it possible to reduce the duration of the contagion (D), the barrier gestures will influence the contagiousness (C), the containment reduces the number of contacts (N) and the vaccination will act on the prevalence of immunized people (P). These mechanisms were clearly in the minds of the governments that imposed these measures. However, as far as we can ascertain, there has been little or no discussion of the expected consequences of the actions taken with respect to these simple formulae.

Some of the concrete questions that decision makers ask during an epidemic include the following. Should demonstrations be banned? If so, from what size? Should school classes, restaurants, and certain businesses be closed? In high schools or universities, what percentage of student participation should be allowed? To answer this type of questions, it would be necessary to stratify the contexts of contamination, and to consider a finer formula for the reproduction number:

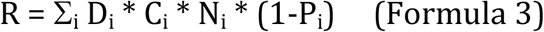

This formula expresses the obvious fact that the total number of contaminations due to a carrier is obtained by the sum of those occurring in all possible (and mutually exclusive) contexts where that carrier has a chance to infect someone. In Formula 3, a possibly different duration D_i_, contagiousness C_i_, number of contacts N_i_ and prevalence of immunized people P_i_ is allowed in each population stratum i. Strata could include for example mass manifestations, public transports, schools, work, shops, leisure activities, homes for the elderly and households. Importantly, each individual could contaminate people in more than one stratum, depending on his/her activities. Again, we are considering a stratification of contaminations, not of individuals, so our terminology may differ somewhat from its traditional meaning in epidemiology.

To use Formula 3 in practice, it is necessary to simplify it. We could for example assume that the duration of the contagion and the prevalence of immunized people is the same in all strata. This is not always possible though. For instance, the prevalence of immunized people will increase more quickly in contaminated strata, such as homes for the elderly, than in the general population. In fact, we shall not consider below the (important but peculiar) stratum of the homes for the elderly, which actually necessitates separate actions and discussions. On the other hand, it is probably fair to assume that contagiousness will be lower for outdoor than for indoor activities, being close to zero and almost negligible for the former. Finally, the number of contacts will strongly depend on the strata, a carrier obviously meeting more people in a crowd or in a public transport than within his/her household.

In this paper, we consider the following simplified formula (or model) to express the reproduction number:

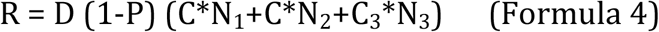

It is a model with three strata and seven parameters. These three strata represent respectively *indoor mass events* (in a closed space with at least 100 persons, e.g. concerts, cinemas, bars, dancing, choirs), *public indoor activities* (including public transport, schools, work, shopping or leisure) and *households* (i.e. families), thought to be three main contexts where contaminations occur. We consider that the number of contacts might be different in each stratum, and assume that contagiousness is identical in the first two strata, but might be higher within households, where the contacts are closer. On the basis of such a simplification of reality, it becomes possible to partly answer some of the questions above, to quantify approximately what the consequences of a government’s measures will be, and to suggest appropriate decisions, as we shall see in the next section.

## Results

In what follows, we examine 20 scenarios involving different parameter values in Formula 4. Results are summarized in Table 1.

**Table 1:**
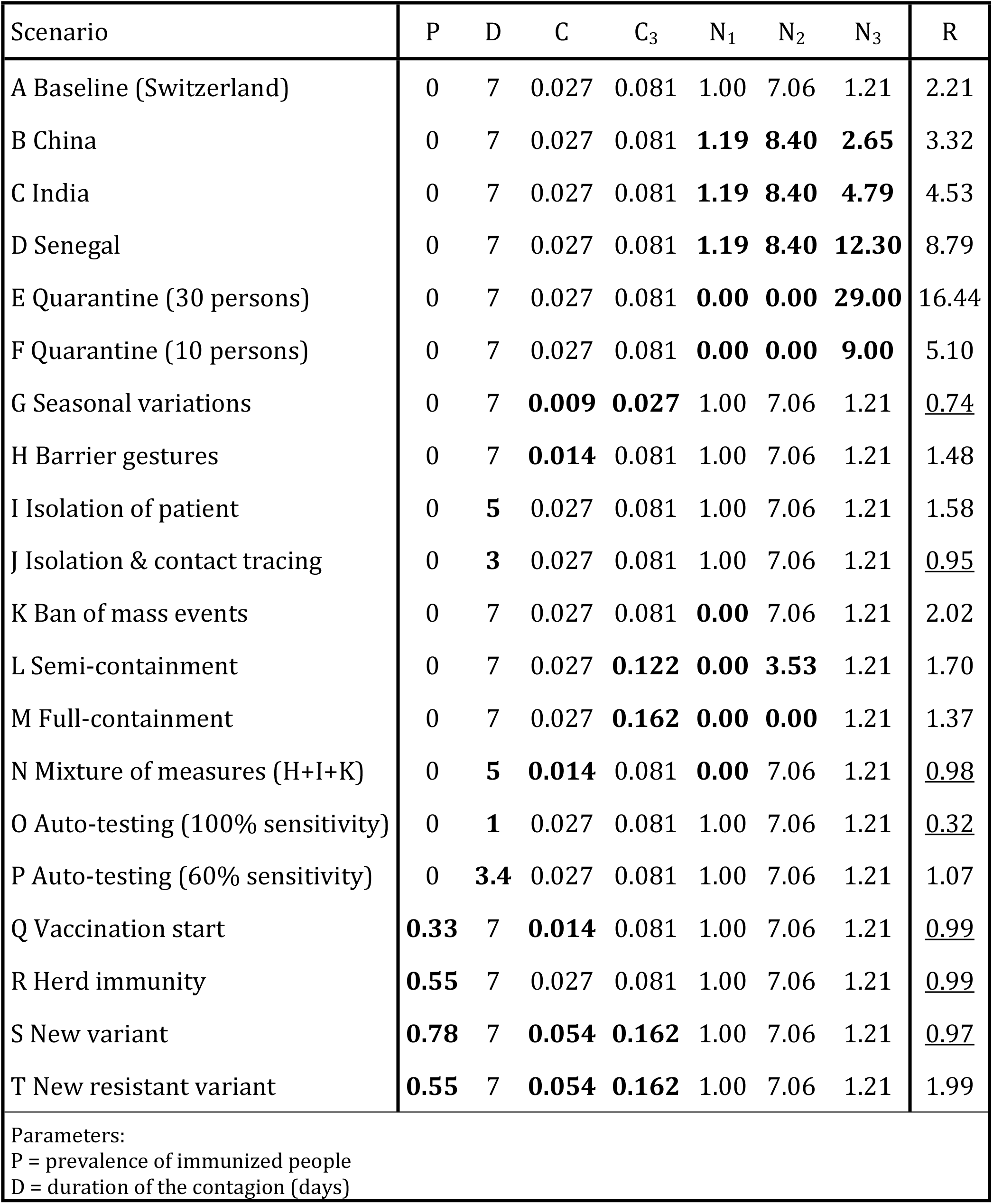
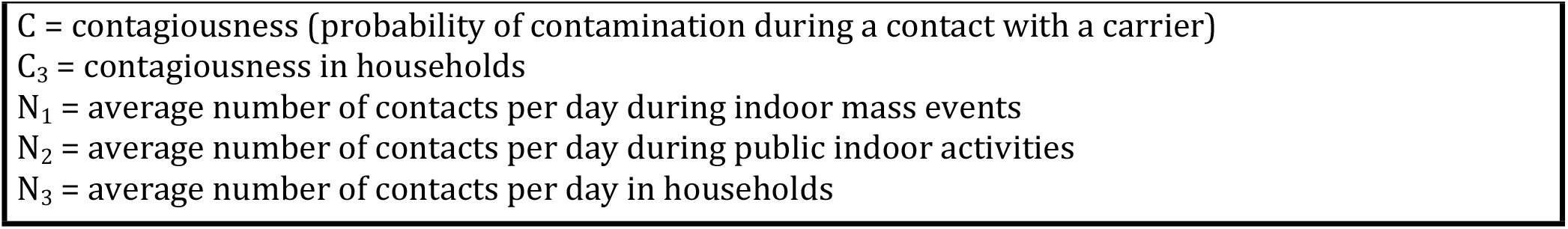
Values of the seven parameters of our model defined by Formula 4 according to 20 scenarios to manage the SARS-Cov-2 epidemic, together with the resulting reproduction number R characterizing the spread of the disease. In bold the parameter values that are different from the baseline scenario (Scenario A). Underlined, the R-values that are less than one.

| Scenario | P | D | C | C<sub>3</sub> | N<sub>1</sub> | N<sub>2</sub> | N<sub>3</sub> | R |
| --- | --- | --- | --- | --- | --- | --- | --- | --- |
| A Baseline (Switzerland) | 0 | 7 | 0.027 | 0.081 | 1.00 | 7.06 | 1.21 | 2.21 |
| B China | 0 | 7 | 0.027 | 0.081 | **1.19** | **8.40** | **2.65** | 3.32 |
| C India | 0 | 7 | 0.027 | 0.081 | **1.19** | **8.40** | **4.79** | 4.53 |
| D Senegal | 0 | 7 | 0.027 | 0.081 | **1.19** | **8.40** | **12.30** | 8.79 |
| E Quarantine (30 persons) | 0 | 7 | 0.027 | 0.081 | **0.00** | **0.00** | **29.00** | 16.44 |
| F Quarantine (10 persons) | 0 | 7 | 0.027 | 0.081 | **0.00** | **0.00** | **9.00** | 5.10 |
| G Seasonal variations | 0 | 7 | **0.009** | **0.027** | 1.00 | 7.06 | 1.21 | <u>0.74</u> |
| H Barrier gestures | 0 | 7 | **0.014** | 0.081 | 1.00 | 7.06 | 1.21 | 1.48 |
| I Isolation of patient | 0 | **5** | 0.027 | 0.081 | 1.00 | 7.06 | 1.21 | 1.58 |
| J Isolation & contact tracing | 0 | **3** | 0.027 | 0.081 | 1.00 | 7.06 | 1.21 | <u>0.95</u> |
| K Ban of mass events | 0 | 7 | 0.027 | 0.081 | **0.00** | 7.06 | 1.21 | 2.02 |
| L Semi-containment | 0 | 7 | 0.027 | **0.122** | **0.00** | **3.53** | 1.21 | 1.70 |
| M Full-containment | 0 | 7 | 0.027 | **0.162** | **0.00** | **0.00** | 1.21 | 1.37 |
| N Mixture of measures (H+I+K) | 0 | **5** | **0.014** | 0.081 | **0.00** | 7.06 | 1.21 | <u>0.98</u> |
| O Auto-testing (100% sensitivity) | 0 | **1** | 0.027 | 0.081 | 1.00 | 7.06 | 1.21 | <u>0.32</u> |
| P Auto-testing (60% sensitivity) | 0 | **3.4** | 0.027 | 0.081 | 1.00 | 7.06 | 1.21 | 1.07 |
| Q Vaccination start | **0.33** | 7 | **0.014** | 0.081 | 1.00 | 7.06 | 1.21 | <u>0.99</u> |
| R Herd immunity | **0.55** | 7 | 0.027 | 0.081 | 1.00 | 7.06 | 1.21 | <u>0.99</u> |
| S New variant | **0.78** | 7 | **0.054** | **0.162** | 1.00 | 7.06 | 1.21 | <u>0.97</u> |
| T New resistant variant | **0.55** | 7 | **0.054** | **0.162** | 1.00 | 7.06 | 1.21 | 1.99 |
| Parameters:<br>P = prevalence of immunized people<br>D = duration of the contagion (days) |  |  |  |  |  |  |  |  |

$C$ = contagiousness (probability of contamination during a contact with a carrier) $C_3$ = contagiousness in households $N_1$ = average number of contacts per day during indoor mass events $N_2$ = average number of contacts per day during public indoor activities $N_3$ = average number of contacts per day in households

### Baseline scenario (onset of the SARS-Cov-2 epidemic in Switzerland)

Since we have 7 parameters in our model, we need 7 information to estimate them. We attempt here to estimate these quantities in a country like Switzerland, where the present authors are living, but this may also be valid in other European countries. For the SARS-Cov-2 epidemic emerging in Wuhan (China), it was reported that the contagion lasted on average D=7 days (two days before the onset of symptoms [15] plus five days after [16]). A similar value of D should probably apply in other parts of the world. At the start of the epidemic, the proportion of immunized people is obviously P=0, while an estimated R=R_0_=2.21 has been reported for Western Europe [17]. On the other hand, a study in Switzerland [18] estimated that D*C_3_=57% of the people sharing the household of a carrier were in turn contaminated, this percentage decreasing to D*C=19% for the contacts of a carrier outside the household. This suggests daily contagiousness values of C_3_=0.57/7=0.081 and C=0.19/7=0.027. The number of people per household in Switzerland is 2.21 [19], meaning that a carrier shared his/her household with an average of N_3_=1.21 persons. To get a seventh information, we assume that 1% of the Swiss population participates to a mass manifestation averaging 100 persons each day, yielding an average number of daily contacts in a mass manifestation of N_1_=0.01*100=1. Clearly, a more precise estimation of this quantity could/should be obtained via a dedicated survey, although this would not fundamentally change the nature of our results. From there, one can estimate an average number of daily contacts outside household of N_2_=(R_0_-D*C*N_1_-D*C_3_*N_3_)/(D*C)=7.06. These are contacts likely to produce contamination, often estimated by fifteen minutes at least of direct or close physical contact [20]. Of note, the R_0_=2.21 contaminations due to a carrier can be decomposed into D*C*N_1_=0.19 (9%) contaminations in the first stratum, D*C*N_2_=1.33 (60%) in the second stratum and D*C_3_*N_3_=0.69 (31%) in the third stratum. Surveys could be used to check whether this decomposition reasonably matches the reality or not, and in the latter case, suggest some appropriate update of the values of parameters in our model.

These data represent Scenario A in Table 1, referred to as the baseline scenario. Without explicit mention of a change, these parameter values of our model will be taken identical in the other scenarios presented below.

### Other places in the world

In China, a meta-analysis provided an estimated value of R_0_=3.32 [21]. Keeping the same contagiousness values as in the baseline scenario, and knowing that the number of people per household is 3.65 in the Hubei region (where it all began) [22], yielding N_3_=2.65, this value of R_0_ would be compatible with an increase of 19% of the contacts in the first two strata compared to the baseline scenario (Scenario B).

This reminds and illustrates that a disease with a given contagiousness and duration of contagion may reach different values of R_0_ depending on the number of contacts and size of the households. As further examples, keeping the data of Scenario B, but considering households of size 5.79 (as in India, Scenario C) or 13.3 (as in Senegal, Scenario D), one would obtain values of respectively R_0_=4.53 and R_0_=8.79, considerably higher than what has been reported in China. Other factors would probably affect the different parameters of our model, for instance climatic factors or the social organization, but these simple calculations show that the reproduction number can vary considerably from one country to another, only depending on the size of the households.

### Quarantines

A strict quarantine strategy is effective in protecting the global population from the arrival of contagious people. However, there are significant risks to those quarantined, as illustrated by the example of the Diamond Princess, where 697 out of 3711 passengers were infected (and 7 died) because of a single carrier at the start of the quarantine [23]. In places less spacious than a ship, quarantined persons could be seen as forming a large household, without any contact outside the household (i.e. with N_1_=N_2_=0). Considering that 30 people are quarantined (i.e. N_3_=29), which could be a school classroom, we end up with a very high reproduction number (R=16.44, Scenario E), such that many of those people will be quickly infected. The calculation also shows that if we would quarantine these people by forming three sub-groups of 10 persons (i.e. N_3_=9), the risks would be lower (R=5.10, Scenario F). Note that these values are just indicative of the intensity of the contagiousness of the disease at the beginning of the quarantine. This does not imply that each carrier will really infect an average of 16.44, respectively of 5.10 other persons, since the prevalence of immunized persons P would quickly increase in such a context.

### Seasonal variations

The onset of the summer 2020 in Europe correlated with a marked decline in the reproduction rate. Assuming a reduction of two thirds of contagiousness (C and C_3_) during the summer would drop the reproduction number below the critical value of 1 without taking any protective measure (R=0.74, Scenario G). Note that the decrease of contagiousness in summer could also be due to the decrease of the intensity of indoor contacts, as people would spend more time outside.

### Protective measures

Apart from a few countries that have implemented a strict containment policy, most nations have had to deal with the pandemic by taking partial protective measures. Formula 4 makes it possible to estimate the potential effectiveness of the different measures. For instance, a realistic application of barrier gestures in public (including wearing a mask), aiming at reducing contagiousness C by half (for example), would not be sufficiently effective on its own, allowing a reduction from R= 2.21 (baseline scenario) to R=1.48 (Scenario H).

A systematic isolation of patients as soon as they show symptoms would typically reduce the average duration of contagion, e.g. from D=7 to D=5 days, to reach R=1.58 (Scenario I). On the other hand, isolating not only the patients but also his/her contacts via efficient (although administratively demanding) contact tracing should make it possible to further reduce this average duration, e.g. to D=3 days, yielding R=0.95 (Scenario J), thus falling below the critical value of 1. The values of D=5 and D=3 days used here are just rough estimations to provide an idea of what could be achieved. Updated and more accurate values of D achievable in these cases could be easily incorporated into Formula 4 if available.

One of the first obvious measure taken by the governments was a total ban on mass events, which in our model is characterized by N_1_=0, allowing for a modest reduction of the reproduction number, to reach R=2.02 (Scenario K). A semi-containment will further reduce the number of public contacts in our model (N_2_), but probably also increase the intensity of contacts among people living in the same household (via more time spent together), and thus intra-household contagiousness (C_3_). Assuming a reduction of 50% of N_2_ (while further assuming N_1_=0) and an increase of 50% of C_3_, we obtain R=1.70 (scenario L), which is not sufficient to fall below the value of 1. Full-containment consist in completely banning any public contact (N_1_=N_2_=0). If, as a result, intra-household contagiousness is doubled compared to the baseline scenario (C_3_=0.162), this yields R=1.37 (Scenario M), which would still be high. Of note, as with quarantines, this would only be an indication of the intensity of contagiousness of the disease at the onset of a full-containment. In theory, a strict full-containment would stop the epidemics, as the proportion of immunized persons living in an infected household would quickly increase, this being not captured by our simple model.

On the other hand, a mixture of measures relatively easy to implement, combining barriers gestures (Scenario H), isolation of patients (Scenario I) and ban of mass events (Scenario K), would allow to formally stabilize the epidemics (R=0.98, Scenario N).

Another promising measure has been the use of auto-tests, which are inexpensive, carried out at home and could significantly reduce the time to contamination if the detected cases are isolated [24]. With tests detecting all cases (100% of sensitivity) already during the asymptomatic stage (e.g. reaching an average duration of contagion of D=1 day, half of the time before the onset of symptoms), we would obtain a low R=0.32 (Scenario O). Unfortunately, such tests may have a modest sensitivity depending on the gold standard (e.g. the number of cycle thresholds used with a PCR test as gold standard) [25]. For example, with 60% of sensitivity, the average duration of contagion would increase to D=0.6*1+0.4*7=3.4 days (since only 60% of cases could be isolated one day after infection, the 40% missed cases remaining contagious during an average of 7 days in this scenario), yielding R=1.07 (scenario P), thus narrowly missing stabilizing the epidemic without additional measures.

### Herd immunity

Formula 4 is also of interest during the last phase of an epidemic, when it comes to estimating how to mitigate the measures, as the prevalence of immunized persons P in the population is increasing (via vaccination or previous infection with the virus).

Shortly after the start of vaccination, when herd immunity reaches, say, P=0.33, it would be sufficient to apply barrier gestures (scenario H) to contain the epidemics, reaching R=0.99 (Scenario Q). In order to avoid any protective measure, herd immunity should reach in our case P=1-1/2.21=0.55 to stabilize the epidemics (R=0.99, Scenario R).

Unfortunately, many viruses tend to develop new variants, with a possible significant change in the contagiousness C and C_3_ of the virus. Recently, searchers estimated a (pre-vaccination) R_0_ value around 5 with the delta variant of SARS-Cov-2 [26, 27] such that contagiousness is roughly doubled compared to our baseline scenario. In such condition, vaccinating 78% of the whole population would be necessary to stop the epidemics (R=0.97, Scenario S). If such a variant were to be partially resistant to vaccination, the reproduction number would increase again. Assuming that only 70% of those immune to the original variant would still be immune to the new one, which would give P=0.78*0.7=0.55, and without any protective measures, we would (almost) be back to the beginning of the epidemics (R=1.99, Scenario T).

## Discussion

In this paper, we proposed a simple model with three strata and seven parameters to evaluate the reproduction number characterizing the spread of an infectious disease. The three strata included in our model are indoor mass events, public indoor activities and households, representing three main contexts where one can be infected. We suggest using such a model to help manage a pandemic during its different phases. It should allow for an expert estimation of the expected effects of the various protective measures taken, an open communication with the public on the assumptions made by the experts in this regard, a continuous assessment of the results through epidemiological surveillance, and finally for suggesting and supporting the gradual release of measures towards the end of the pandemic. In case of discrepancies between what has been assumed and what has been observed in reality, some assumptions could be updated and the model re-evaluated using different parameter values.

To estimate the seven parameters in our model, seven assumptions or pieces of information are needed. Some is not be too difficult to obtain, such as the average household size in a given country. Some could be borrowed from other countries, such as the average duration of contagion of the disease. Others could be obtained through specific surveys, such as the frequency of population participation in a mass event, the proportion of infections occurring within a household, or by investigating the sources of contamination [28]. Finally, some information should simply be guessed or assumed on the basis of the scientific literature, with the possibility (and duty) of being updated to become more and more accurate along the course of the pandemic.

In the Results section, we have illustrated how such information could be gathered and the model used to manage the SARS-Cov2-epidemic, considering as baseline scenario the onset of the epidemic in Switzerland (or in a European context), although it could easily be recalculated to fit other configurations. Although our model is simple, we could attempt to evaluate the influence of most of the potential protective measures that could be applied by governments, either alone or simultaneously.

Using debatable but transparent assumptions, we were able to identify some scenarios that would allow us to reach a reproduction number of less than one, a necessary condition for the stabilization of an epidemic. One of the most effective scenario would be just to rely on seasonal variations, as a virus is generally less contagious in summer than in winter. Of the measures we could act on, simple isolation of patients might not be effective enough, while isolation and contact tracing might be sufficient, but cumbersome to implement. Semi-containment, accompanied by a ban on mass events and the application of gesture barriers might be a more pragmatic alternative. The best approach would probably be a systematic application of rapid auto-tests with isolation of the detected cases. Both interventions are inexpensive and do not cause too many social restrictions. The possibly low sensitivity of these tests will, however, mitigates the effects of this attractive measure, although it will remain useful, if not to stop the pandemics, at least to identify clusters, for example. When available, vaccination of a significant proportion of the population is another effective measure, unless a resistant variant of the virus emerges. Of course, all these examples are theoretical and, as already mentioned, each parameter value we have used can be criticized and modified to obtain possibly more accurate evaluations.

An important aspect of our model is to consider a specific stratum for households. The focus on stratifying for household contagion is not new [29, 30]. Surprisingly, almost no action has been recommended to reduce this factor during the pandemic. De facto, many professionals separated their spouses during the acute phases of the pandemic, especially when masks and other protective measures were lacking. This is likely to have had an impact. The effect of such an action at the level of the population could be investigated using our model.

Many authors have compared the dangerousness of the coronavirus with other viruses, such as influenza for example, based on R_0_ values. Our model helps to better understand and illustrate that an R_0_ can vary considerably depending on the context and that it would be better to analyze contagiousness directly [31]. Even with a much more stable virus like measles, there are large variations in R_0_ estimates that are poorly understood [32]. The SARS-Cov-2 pandemic surprised by its virulence, with an initial underestimation of its dangerousness. The epicenter of the onset of the pandemic was in China, which has a relatively small household size. Our calculations show, that the epidemic would probably have appeared more dangerous if it had originated in India, for example.

Our model has of course limitations. An important one is that we are not able to recognize that a strict full-containment would be effective (even if it is difficult to implement and to get a population to accept), as shown during the first wave of SARS-Cov-2 epidemic in China. This is because the proportion of immunized persons in our model is assumed to be stable and identical in all three strata. In particular, we do not capture in our model that the proportion of immunized persons in an infected household will increase rapidly in the event of a full-containment, so that there will be no one left to infect, thus stopping the epidemics. This will also apply to a lesser extend in a semi-containment [33,34]. We are currently investigating ways to extend our model in this respect, to take into account the fact that herd immunity is reached more quickly in households than in the general population.

Other limitations of our model include the following. We are not able to assess the role of border controls to limit the circulation of virus variants. As mentioned in the Methods section, we ignored the important stratum of the homes for the elderly, while we did not consider the specific situation of children. A model with more than three strata might be preferable, although this would imply a larger number of parameters to estimate.

In the end, our model is probably too simple. Note that the use of more sophisticated models (SEIR compartmental models or others) provided similar results (advantage or early detection and mixture of containments measures) but are much more difficult to implement [35,36]. Scientific studies have also struggled to demonstrate the effectiveness of e.g. semi-containment measures and the effects of climatic factors. This is why we still believe that despite its obvious limitations, our simple and pedagogical model can be extremely useful in helping a government to think, decide and evaluate what to do during a pandemic. Its use will bring more transparency and public support than the use of more sophisticated models, also to manage future pandemics with different characteristics than SARS-Cov-2.

## Data Availability

No data (theoretical research article)

